# Cross-Center Validation of Deep Learning Model for Musculoskeletal Fracture Detection in Radiographic Imaging: A Feasibility Study

**DOI:** 10.1101/2024.01.17.24301244

**Authors:** Robert Hrubý, Daniel Kvak, Jakub Dandár, Anora Atakhanova, Matěj Misař, Daniel Dufek

## Abstract

Fractures, often resulting from trauma, overuse, or osteoporosis, pose diagnostic challenges due to their variable clinical manifestations. To address this, we propose a deep learning-based decision support system to enhance the efficacy of fracture detection in radiographic imaging. For the purpose of our study, we utilized 720 annotated musculoskeletal (MSK) X-rays from the MURA dataset, augmented by bounding box-level annotation, for training the YOLO (You Only Look Once) model. The model’s performance was subsequently tested on two datasets, sampled FracAtlas dataset (Dataset 1, 840 images, *n*_NORMAL_ = 696, *n*_FRACTURE_ = 144) and own internal dataset (Dataset 2, 124 images, *n*_NORMAL_ = 50, *n*_FRACTURE_ = 74), encompassing a diverse range of MSK radiographs. The results showed a Sensitivity (*Se*) of 0.910 (95% CI: 0.852–0.946) and Specificity (*Sp*) of 0.557 (95% CI: 0.520–0.594) on the Dataset 1, and a *Se* of 0.622 (95% CI: 0.508–0.724) and *Sp* of 0.740 (95% CI: 0.604–0.841) on the Dataset 2. This study underscores the promising role of AI in medical imaging, providing a solid foundation for future research and advancements in the field of radiographic diagnostics.

## 1 Introduction

Bone fractures, defined as disruptions in bone continuity, arise from diverse etiologies including trauma, stress injuries, and pathological conditions such as osteoporosis [1]. These fractures, irrespective of the patient’s age, manifest a spectrum of clinical presentations ranging from mild symptoms like pain and swelling to severe complications including deformity and functional impairment of the impacted region [2]. Clinically, fracture diagnosis incorporates an injury history, physical examination, and symptom evaluation, notably pain, edema, morphological alterations, abnormal mobility, and occasionally crepitus – a palpable or audible friction in the fractured bone. However, symptomatology in certain fracture types, such as closed or stress fractures, might be subtle or non-specific [3].

Diagnostic approaches for fracture detection and confirmation predominantly utilize X-ray imaging as the initial modality, offering detailed bone structure visualization and fracture identification [4]. Supplementary imaging techniques, including computed tomography (CT), magnetic resonance imaging (MRI), or ultrasound, are employed for complex cases or specific fracture types where musculoskeletal (MSK) X-rays are inadequate. Fracture management strategies depend on fracture characteristics like type, location, and severity, ranging from non-invasive treatments like splint immobilization to surgical interventions using internal osteosynthesis with screws, pins, wires, or external fixation devices [5]. Rehabilitation, encompassing physical therapy and exercises, is crucial for restoring function and strength to the affected region.

In the realm of clinical decision-making, the integration of deep learning-based decision support software for radiograph interpretation marks a significant advancement. Despite promising developments, existing algorithms for fracture detection face practical limitations, including the inability to analyze all body parts simultaneously or detect multiple fractures in a single X-ray, which are common in clinical practice [6]. Leveraging sophisticated deep learning models, such software enhances fracture detection accuracy on radiographs, alleviates the diagnostic workload of radiologists, and augments patient outcomes through expedited and precise diagnostic processes [7].

## 2 Materials and Methods

### 2.1 Training Data

For this feasibility study, the publicly available MURA dataset, further described in the study “MURA: Large Dataset for Abnormality Detection in Musculoskeletal Radiographs” [8] (Figure 1) was selected to validate the design and development of the proposed deep learning algorithm. As this dataset contains MSK X-rays annotated only at the label-level, the images required to be annotated at the bounding box-level.

**Figure 1:**
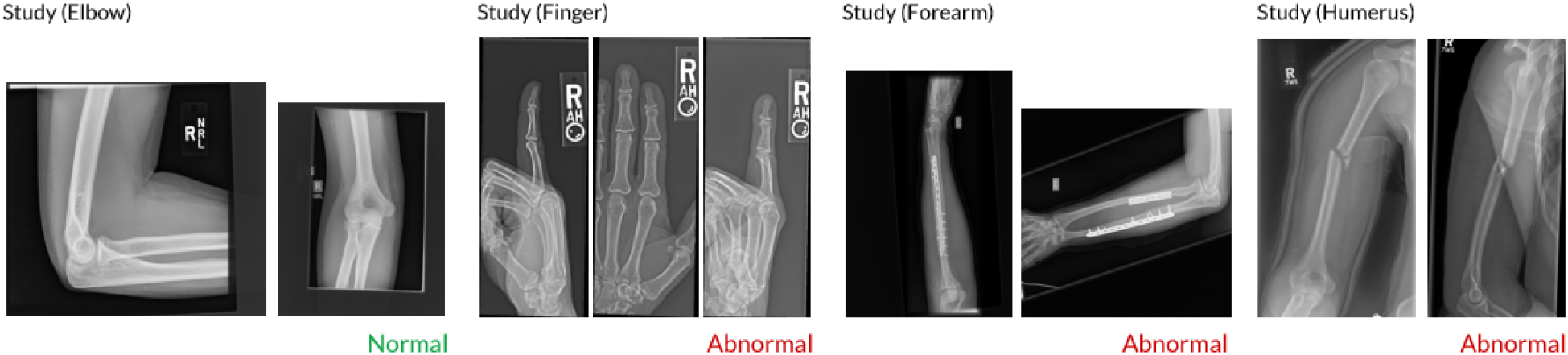
Examples of MSK X-rays of the upper extremity from the MURA dataset, with each study manually labeled by radiologists as normal (absence of fracture) or abnormal (presence of fracture). These examples show a normal study of the elbow, an abnormal study of the finger with degenerative changes, an abnormal study of the forearm demonstrating surgical fixation with plate and screws for radius and ulna fractures, and an abnormal study of the humerus with a fracture.

A total of 720 images with positive fracture findings were randomly extracted from the primary dataset of 40,561 skeletal radiographs (14 863 studies in total). Subsequently, the selected dataset was annotated by an in-house radiologist at the bounding-box labeling level. For training our deep learning model, we utilized 30 images of the shoulder, 249 images of the upper arm, 123 images of the elbow, 117 images of the forearm, 68 images of the wrist, and 133 images of the hand and fingers, containing a total of 832 bounding boxes indicating the presence of a fracture and localization (Table 1).

**Table 1:** Number of training images and bounding boxes for the indication of fracture.

| Anatomic Area of interest | $n$ (images) | $n$ (bounding-boxes) |
| --- | --- | --- |
| Shoulder | 30 | 31 |
| Humerus | 249 | 328 |
| Elbow | 123 | 137 |
| Forearm | 117 | 131 |
| Wrist | 68 | 71 |
| Hand and Fingers | 133 | 134 |
| *Total* | *720* | *832* |

### 2.2 Model Architecture

YOLO (You Only Look Once) [9] (Figure 2, Figure 3) is a single-stage regression model for object detection. This model uniquely combines object localization and classification in a single pass through a convolutional neural network, enabling fast, real-time detection. During analysis, the entire radiographic image is passed through the network, which is crucial for higher detection accuracy in scenarios where contextual information about bone structure is vital.

**Figure 2:**
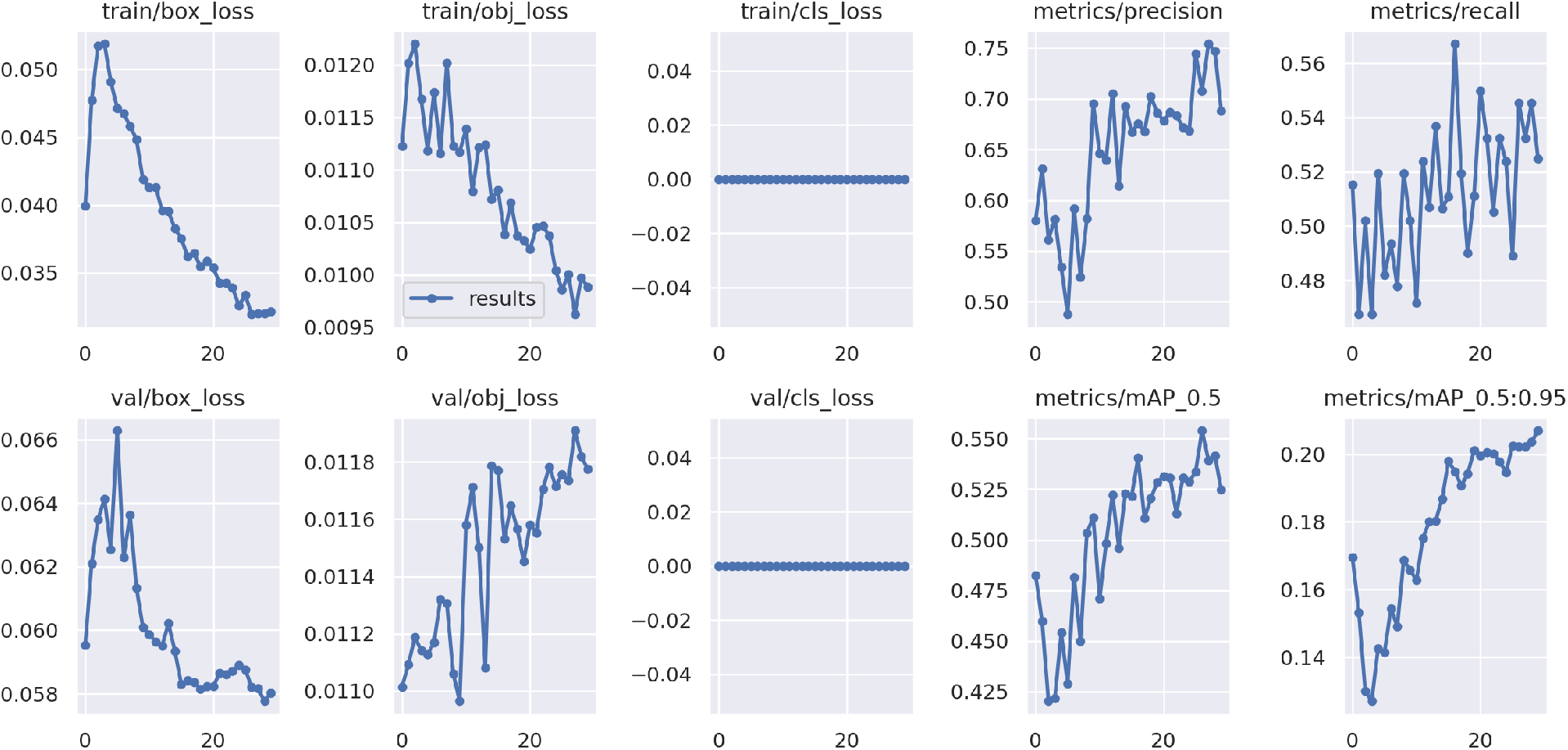
Train/loss and val/loss metrics for the indication of fracture.

**Figure 3:**
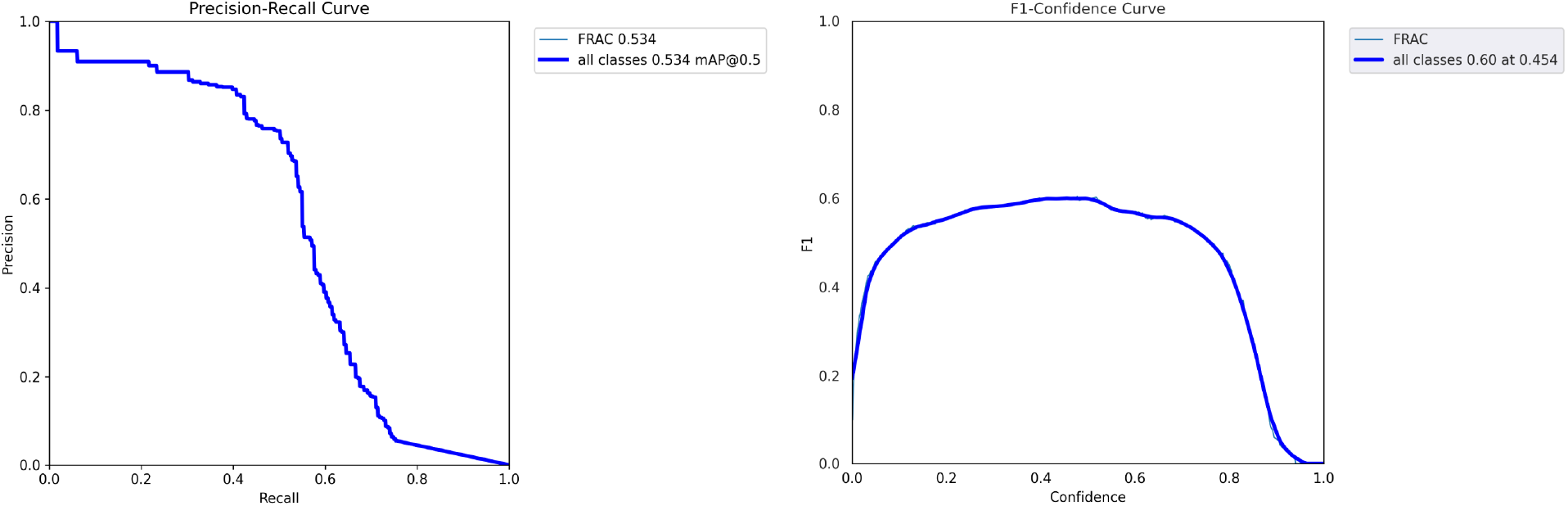
Precision-Recall Curve a F1-Confidence Curve for the indication of fracture.

Developed using the Darknet framework, YOLO is optimized for both CPU and GPU computation, making it suitable for diverse clinical settings. The model outputs bounding boxes, each representing a detected fracture site. The bounding box is defined by:

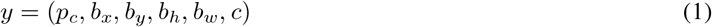

where *p*_*c*_ is the probability of fracture presence, (*b*_*x*_, *b*_*y*_) are the coordinates of the bounding box center, (*b*_*h*_, *b*_*w*_) are the dimensions of the bounding box, and *c* represents the class of the object, in this case, a fracture. The deep learning model’s output is then layered on the original musculoskeletal X-ray image, highlighting the regions of interest with bounding boxes, thereby aiding radiologists in identification of fractures.

### 2.3 Assessment

To evaluate the ability of the deep learning algorithm to evaluate the presence or absence of fractures on MSK X-rays, the proposed model was retrospectively validated on a multicenter dataset, including both external, publicly available sample data, and internal testing data (Table 2). Dataset 1 contains a randomly selected sample from the FracAtlas dataset, described in more detail in the study “FracAtlas: A Dataset for Fracture Classification, Localization, and Segmentation of Musculoskeletal Radiographs” [10], comprising a total of 840 MSK images (*n*_NORMAL_ = 696, *n*_FRACTURE_ = 144). Dataset 2 consists of internal data, annotated by our in-house radiologist, encompassing a total of 124 MSK images radiographs (*n*_NORMAL_ = 50, *n*_FRACTURE_ = 74). Ground truth was established on image-level.

**Table 2:** Test data distribution for Dataset 1 and Dataset 2.

| Dataset | $n$ | $n_{\text{NORMAL}}$ | $n_{\text{FRACTURE}}$ |
| --- | --- | --- | --- |
| Dataset 1 | 840 | 696 | 144 |
| Dataset 2 | 124 | 50 | 74 |

### 2.4 Statistical Analysis

The clinical performance of the proposed deep learning algorithm was validated using key statistical metrics. Sensitivity (*Se*) refers to the algorithm’s ability to correctly identify cases where a fracture is present on musculoskeletal (MSK) X-ray images. Specificity ((*Sp*) refers to the algorithm’s ability to correctly identify that a fracture is not present in individuals without such a condition. The Wilson score interval was used to calculate the 95% confidence intervals (CIs) for both *Se* and *Sp*, ensuring an accurate measure of the algorithm’s diagnostic efficacy.

To further assess the performance of the deep learning algorithm, we use the metrics of positive (*PPV*) and negative predictive value (*NPV*), and positive (*PLR*) and negative likelihood ratio (NLR). *PPV* indicates the probability that a patient with a positive diagnostic result actually has a fracture, while NPV indicates the probability that a patient with a negative result does not show a fracture. These values are directly influenced by the prevalence of the positive finding in the study population. *PLR* and *NLR* provide information on how much the probability of having a fracture increases or decreases depending on the test result. The *PLR* indicates the increase in the probability of a fracture associated with a positive result, while the *NLR* indicates how the probability decreases in the event of a negative result. To provide a more accurate estimate of the reliability of these metrics, we calculated 95% CIs. For *PPV* and *NPV*, we used binomial approximations for CIs based on the standard error for a binomial distribution. For *PLR* and *NLR*, the intervals were calculated by log transforming the values to include potential variability in the data.

## 3 Results

The proposed deep learning algorithm correctly predicted 131 images (TP) containing one or more fractures out of a total of 144 positive skeletal X-ray images for Dataset 1, while it misclassified 308 negative images (FP) out of a total of 696 images. For Dataset 2, the algorithm correctly predicted 46 images (TP) out of a total of 74 positive examples, while incorrectly classifying 13 (FP) images without abnormality (Figure 4).

**Figure 4:**
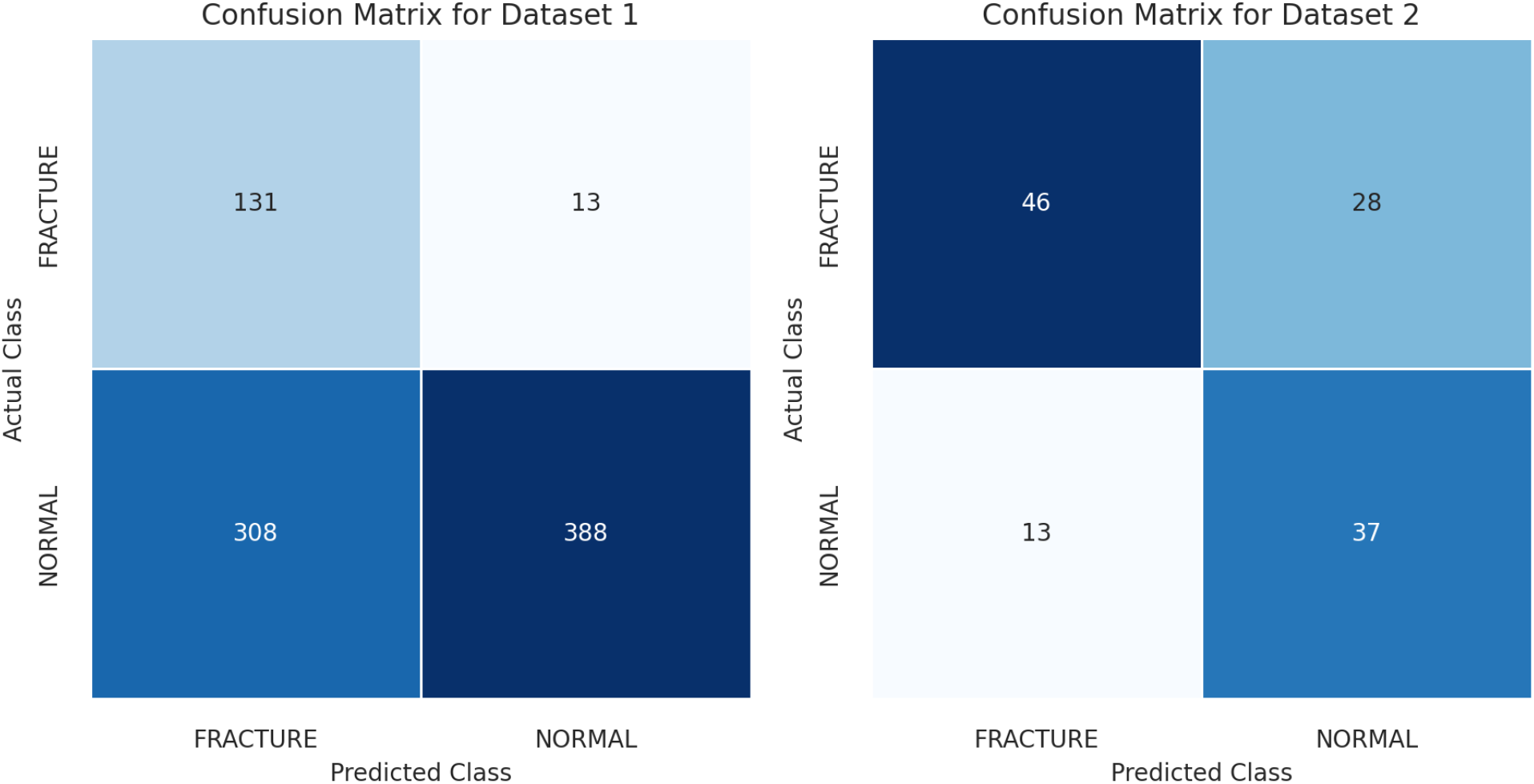
Confusion matrix illustrating the performance of the proposed deep learning algorithm.

**Figure 5:**
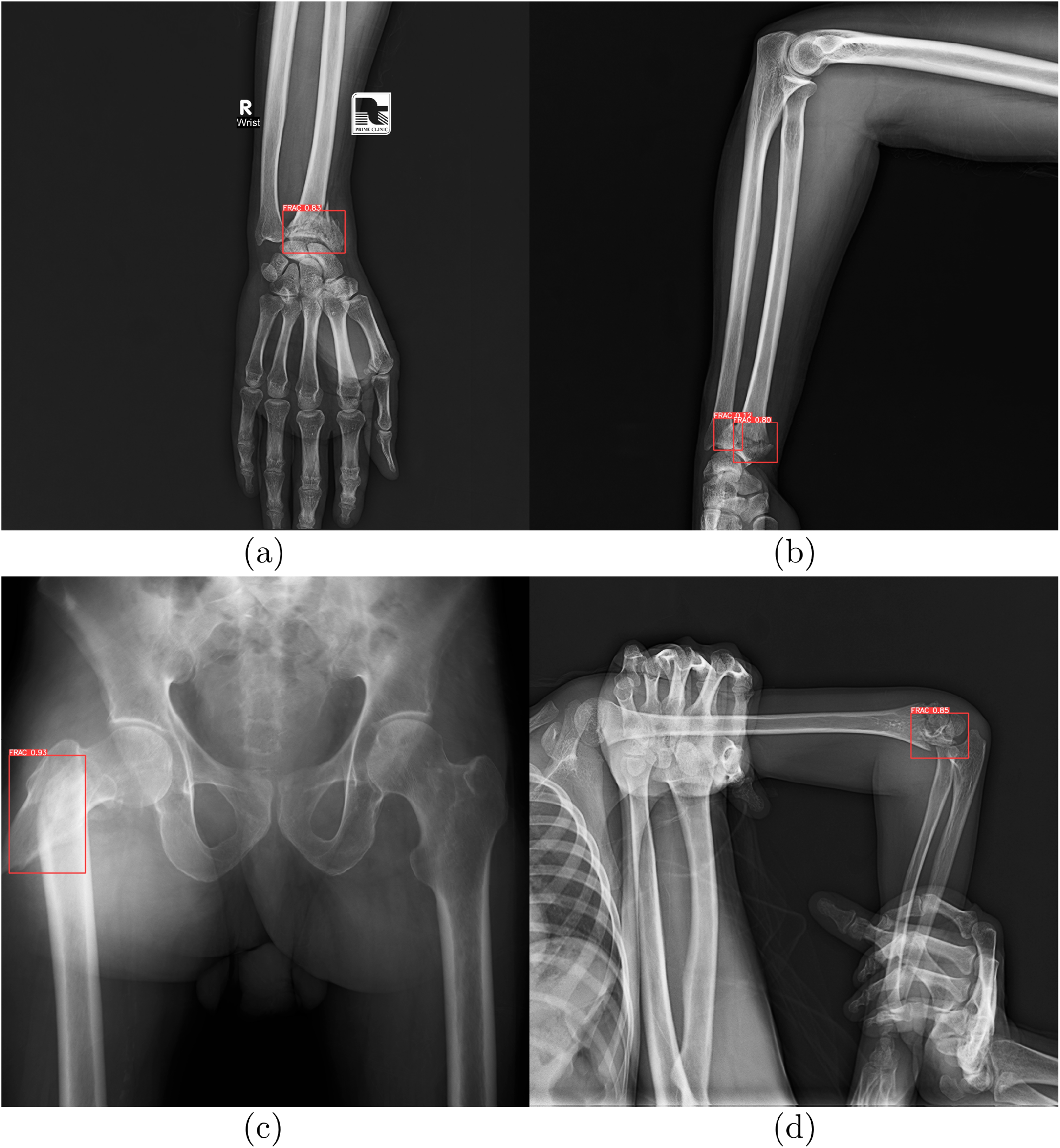
Example predictions of the proposed deep learning algorithm trained to identify skeletal fractures in X-ray images. The examples show the true positive (TP) predictions of the algorithm on the input images from Dataset 1: (a) frontal image of the hand and wrist, (b) lateral image of the forearm, (c) frontal image of the hip and pelvis, (d) lateral image of the elbow.

**Figure 6:**
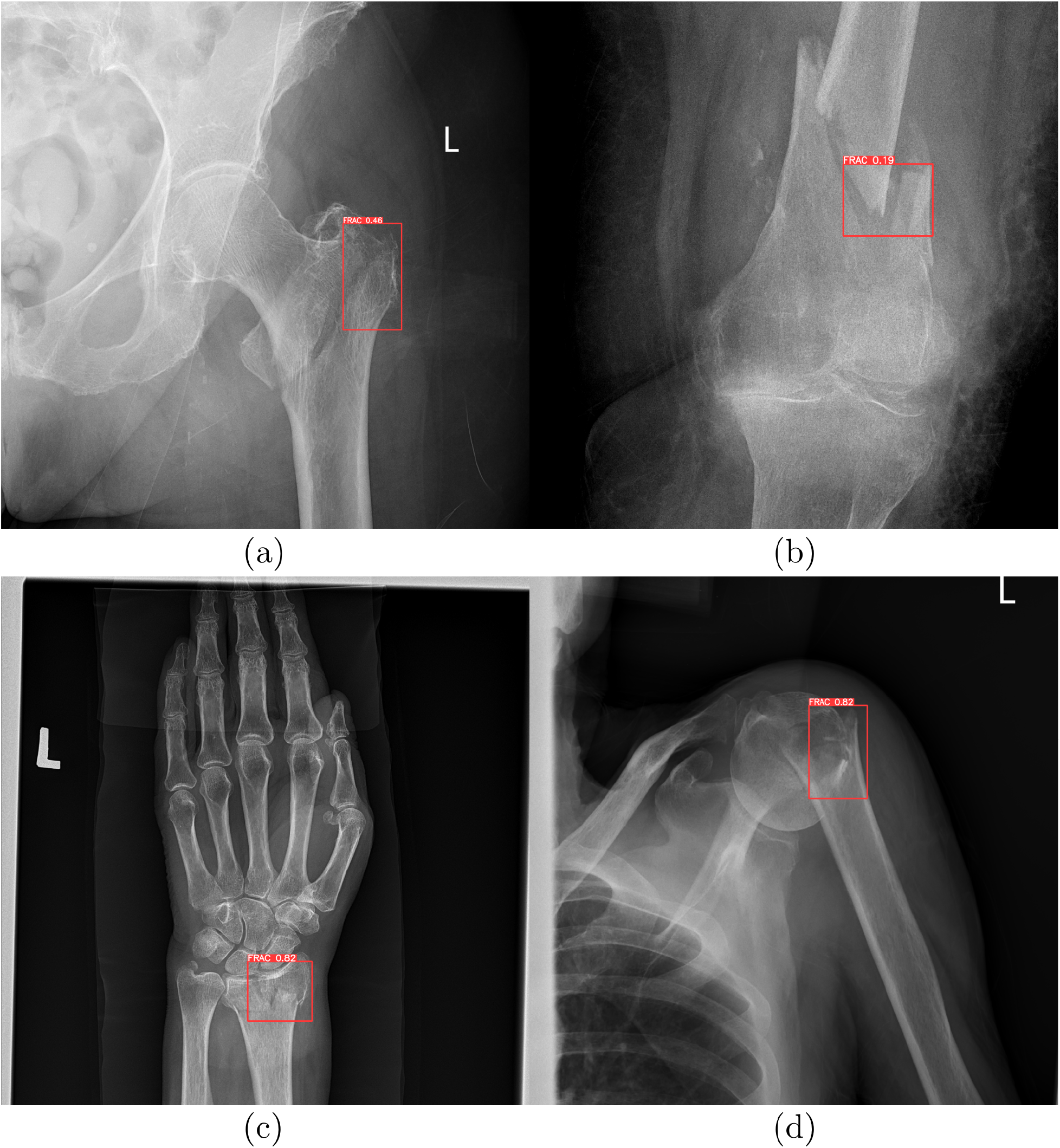
Example predictions of the proposed deep learning algorithm trained to identify skeletal fractures in X-ray images. The examples show the true positive (TP) predictions of the algorithm on the input images from Dataset 2: (a) frontal image of the hip, (b) frontal image of the knee, (c) frontal image of the wrist, (d) frontal image of the shoulder.

This corresponds to a sensitivity (Se) of 0.910 (95% CI: 0.852-0.946) and specificity (Sp) of 0.557 (95% CI: 0.520-0.594) on Dataset 1, and Se of 0.622 (95% CI: 0.508-0.724) and Sp of 0.740 (95% CI: 0.604-0.841) on Dataset 2 (Table 3).

**Table 3:**
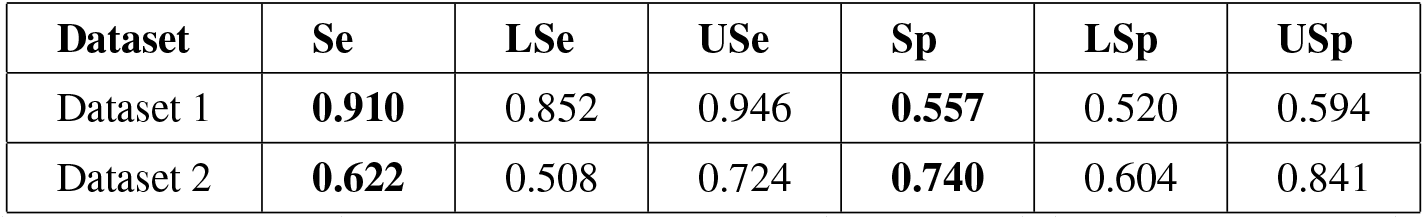
Pooled sensitivity (Se) and specificity (Sp) results for the proposed deep learning algorithm, including the corresponding 95% Confidence Interval (CI) estimates.

The algorithm’s performance was further assessed using Positive Predictive Value (PPV) and Negative Predictive Value (NPV), which respectively indicate the probability that patients with positive and negative test results truly have and do not have the condition. For Dataset 1, the PPV was 0.298 (95% CI: 0.256-0.341) and NPV was 0.968 (95% CI: 0.950-0.985). For Dataset 2, the PPV was 0.780 (95% CI: 0.674-0.885) and NPV was 0.569 (95% CI: 0.449-0.690). Likelihood Ratios (LRs), which combine sensitivity and specificity, were also calculated. The Positive Likelihood Ratio (PLR) quantifies how much the odds of the disease increase when a test is positive, while the Negative Likelihood Ratio (NLR) indicates how much the odds of the disease decrease when a test is negative. For Dataset 1, the PLR was 2.056 (95% CI: 2.008-2.968) and NLR was 0.162 (95% CI: 0.140-0.187). For Dataset 2, the PLR was 2.391 (95% CI: 1.926-2.968) and NLR was 0.511 (95% CI: 0.447-0.585) (Table 4.

**Table 4:** Pooled positive (PPV) and negative predictive value (NPV), and positive (PLR) and negative likelihood ratio (NLR) for the proposed deep learning algorithm, including the corresponding 95% Confidence Interval (CI) estimates.

| Dataset | PPV | LPPV | UPPV | NPV | LNPV | UNPV |
| --- | --- | --- | --- | --- | --- | --- |
| Dataset 1 | **0.298** | 0.256 | 0.341 | **0.968** | 0.950 | 0.985 |
| Dataset 2 | **0.780** | 0.674 | 0.885 | **0.569** | 0.449 | 0.690 |
| Dataset | PLR | LPLR | UPLR | NLR | LNLR | UNLR |
| Dataset 1 | **2.056** | 2.008 | 2.968 | **0.162** | 0.140 | 0.187 |
| Dataset 2 | **2.391** | 1.926 | 2.968 | **0.511** | 0.447 | 0.585 |

These values suggest that while the algorithm is highly effective in correctly ruling out fractures (high NPV), its ability to confirm the presence of fractures (PPV) varies significantly between datasets. The LRs indicate a moderate increase in the probability of a fracture being present with a positive test result, and a moderate to significant decrease in probability with a negative test result.

## 4 Discussion

The study’s findings highlight the deep learning algorithm’s proficiency in detecting musculoskeletal (MSK) fractures in X-ray images. The algorithm exhibited high sensitivity (*Se*) in Dataset 1, indicating its strong capability in correctly identifying fractures. However, the lower *Se* observed in Dataset 2 suggests potential limitations in diverse clinical scenarios or variations in fracture types and imaging qualities. Specificity (*Sp*) varied significantly between the two datasets, with a notably lower *Sp* in Dataset 1 compared to Dataset 2. This variation may reflect differences in the datasets’ characteristics, such as the distribution of fractures or differences in X-ray interpretation. These findings underscore the importance of considering dataset diversity when assessing algorithm performance in a clinical context.

### 4.1 Limitations

The high negative predicitive value (*NPV*) across both datasets reinforces the algorithm’s effectiveness in ruling out fractures, which is critical in clinical decision-making. However, the variability in positive predictive value (*PPV*) points to the need for cautious interpretation of positive results and suggests that the algorithm’s role should initially be as a supportive tool rather than a definitive diagnostic solution.

## 5 Conclusion

In conclusion, this study validates the potential of the YOLO-based deep learning algorithm for fracture detection in MSK radiography. Its high sensitivity in some cases and consistently high negative predicitive value are promising, but the variability in specificity and positive predictive value indicates the need for further optimization and broad-scale validation. These findings advocate for the integration of such AI tools in radiographic analysis, complementing the expertise of radiologists to enhance diagnostic accuracy and improve patient care outcomes. Future work should focus on collecting extensive training dataset with including ground truth, refining the algorithm, and validating its performance in diverse, real-world clinical settings.

## Data Availability Statement

This study uses MURA dataset, presented in “MURA: Large Dataset for Abnormality Detection in Musculoskeletal Radiographs“, FracAtlas dataset, presented in “FracAtlas: A Dataset for Fracture Classification, Localization and Segmentation of Musculoskeletal Radiographs“, and internal dataset from Carebot s.r.o., which is available upon written request. This study was enabled by contract for the transfer of X-ray images for medical research purposes (Smlouva o předání skiagrafických snímku° pro účely medicínského výzkumu) between Nemocnice Šumperk, a.s. and Carebot s.r.o., signed on 14 December 2023 in Prague, Czech Republic. The Ethics Committee of Nemocnice Šumperk, a.s approved the transfer of patient studies for the research project. Patient consent was waived due to Regulation 2016/679 of the European Parliament and of the Council of 27 April 2016 on the protection of natural persons with regard to the processing of personal data and on the free movement of such data, and the repealing of Directive 95/46/EC (General Data Protection Regulation); i.e., the images were stripped of all direct or indirect identifiers without the possibility of retrospective patient identification.

